# Effects of Mindfulness-Based Interventions on Executive Function in Children and Adolescents: A Systematic Review and Meta-Analysis

**DOI:** 10.64898/2026.04.18.26351184

**Authors:** Nathan Mingyang Li

## Abstract

**Background:** Mindfulness-based interventions (MBIs) have been increasingly adopted in educational settings to support cognitive development in youth. Executive function (EF)—encompassing inhibitory control, working memory, and cognitive flexibility—is a plausible target of MBI given its reliance on attention regulation. However, prior reviews have yielded mixed conclusions, partly due to inconsistent construct definitions and the pooling of heterogeneous outcome measures.

**Objectives:** To (1) estimate the pooled effect of MBI on EF in youth aged 3–18 years using only construct-validated, direct EF measures, (2) examine potential moderators including age group, EF domain, and risk of bias, and (3) test dose-response relationships via meta-regression on intervention duration.

**Methods:** We searched PubMed, PsycINFO, CINAHL, Scopus, and Web of Science from inception to March 2026, supplemented by reference-list searches from two existing systematic reviews and a scoping review. Only English-language publications were eligible. Eligible studies were randomised controlled trials (RCTs) or quasi-RCTs of MBI (excluding yoga-only interventions) in typically developing youth, with at least one direct behavioural or computerised EF outcome. Risk of bias was assessed using Cochrane RoB 2. Hedges’ *g* was computed for each study, and pooled using a DerSimonian–Laird random-effects model. Subgroup analyses by age group, EF domain, and risk of bias were conducted, alongside leave-one-out sensitivity analyses, Egger’s regression test, trim-and-fill, and Knapp-Hartung-adjusted meta-regression on intervention duration. Evidence certainty was rated using GRADE.

**Results:** Thirteen RCTs (nine school-age, four preschool; total *N* = 1,560) met inclusion criteria. The pooled effect was *g* = 0.365 (95% CI 0.264 to 0.465; *p* < .00001), with negligible heterogeneity (*I*^2^ = 0.0%; *Q* = 6.76, *p* = .87). Effects were consistent across age groups (school-age *g* = 0.389; preschool *g* = 0.318) and EF domains (inhibitory control, working memory, cognitive flexibility; *p*_between_ = .60). Meta-regression on intervention duration (4–20 weeks) was non-significant (*p* = .79). The effect was robust in leave-one-out analyses, in the low risk-of-bias subgroup (*g* = 0.361; *k* = 8), and after trim-and-fill adjustment (*g* = 0.354). The 95% prediction interval (0.252 to 0.477) was entirely positive. GRADE certainty was rated MODERATE, downgraded once for risk of bias.

**Conclusions:** MBIs appear to produce a small, statistically significant improvement in EF in youth aged 3–18 years, with moderate certainty of evidence per the GRADE framework. The effect is consistent across preschool and school-age samples and across EF domains, with no significant dose-response relationship within the 4–20 week range studied. Emerging mediation evidence suggests that EF improvement may serve as an important pathway through which MBI supports emotion regulation, though this requires replication. Further large-scale, pre-registered RCTs with active control conditions and longitudinal follow-up are warranted.

## 1. Introduction

Mindfulness has been defined as paying attention in a particular way: on purpose, in the present moment, and non-judgementally (Kabat-Zinn, 1994). Over the past two decades, mindfulness-based interventions (MBIs) have been adapted for youth in educational, clinical, and community settings, with curricula such as MindUP (Schonert-Reichl & Lawlor, 2010), the Kindness Curriculum (Flook et al., 2015), and Inner Kids (Flook et al., 2010) designed specifically for children and adolescents.

Executive function (EF) refers to a family of top-down cognitive processes that enable goal-directed behaviour in the face of novel or demanding situations (Diamond, 2013). The prevailing framework identifies three core components: inhibitory control (the ability to suppress prepotent responses), working memory (the capacity to hold and manipulate information), and cognitive flexibility (the ability to shift between mental sets or tasks) (Miyake et al., 2000; Diamond, 2013). These core components support higher-order capacities including planning, problem-solving, reasoning, and pragmatic social communication, though the present review focuses on the three foundational components as defined by Diamond (2013) to maintain construct specificity. These components are subserved by prefrontal and anterior cingulate cortical networks that undergo protracted development from early childhood through adolescence (Blakemore & Choudhury, 2006). EF is a robust predictor of academic achievement, social competence, and mental health across development (Blair & Razza, 2007; Moffitt et al., 2011), making it a high-priority target for early intervention.

The theoretical link between mindfulness practice and EF rests on shared neural substrates. Mindfulness meditation engages attention regulation, body awareness, and emotion regulation processes (Tang, Holzel, & Posner, 2015) that overlap with the prefrontal networks supporting EF. In adults, there is evidence suggesting that attention regulation may serve as a foundational mechanism through which MBI exerts its effects: by strengthening attentional control, mindfulness practice may cascade into improvements across multiple cognitive domains, including but not limited to EF (Jha, Krompinger, & Baime, 2007; Posner & Rothbart, 2007). In adults, meta-analytic evidence indicates that MBI improves cognitive performance, including measures of attention and EF (Chiesa, Calati, & Serretti, 2011; Zainal & Newman, 2023). Whether these benefits extend to the developing brain of children and adolescents is a question of considerable practical importance.

Several prior reviews have examined this question with varying conclusions. Zenner, Herrnleben-Kurz, and Walach (2014) found a moderate pooled effect of school-based MBI on cognitive outcomes in youth (*g* = 0.40, *k* = 19), though the cognitive outcome category included diverse measures spanning attention, memory, and academic performance. Mak, Whittingham, Cunnington, and Boyd (2018) conducted a systematic review of 13 RCTs of MBI for attention and EF in children and adolescents, but could only meta-analyse two studies (Stroop test outcomes from yoga interventions), finding no significant effect. Dunning et al. (2019) reported a small pooled effect for cognition in their comprehensive review of 33 youth MBI RCTs (*d* = 0.19, 95% CI 0.05 to 0.33), though their cognition category subsumed measures of attention, EF, and academic performance. Most recently, Kander, Heininga, and de Moor (2024), pooling 32 studies examining mindfulness-based EF interventions in youth, reported *g* = 0.34 (95% CI 0.22 to 0.46).

A notable limitation of the existing literature is the inconsistent operationalisation of EF. Some reviews pool EF with broader cognitive constructs such as sustained attention, processing speed, or academic achievement (Dunning et al., 2019; Zenner et al., 2014), potentially obscuring the specificity of MBI effects on core EF processes. Others include yoga-only interventions (Mak et al., 2018), which may operate through different mechanisms than mindfulness meditation per se. Furthermore, the preschool period—a time of rapid EF development (Zelazo, 2015)—has often been underrepresented in prior reviews, with a recent scoping review identifying 17 eligible preschool RCTs that had not been synthesised in earlier meta-analyses (Author, 2025).

The present review addresses these limitations by applying a strict, construct-validated definition of EF (Diamond, 2013), restricting inclusion to MBI (excluding yoga-only), requiring direct behavioural or computerised EF measures (excluding teacher- or parent-rated behaviour scales that do not specifically assess EF), and spanning the full age range of 3–18 years. We aimed to (1) estimate the pooled effect of MBI on EF in youth, and (2) examine whether this effect is moderated by age group (preschool vs. school-age) or methodological quality.

## 2. Methods

This systematic review was conducted and reported in accordance with the Preferred Reporting Items for Systematic Reviews and Meta-Analyses (PRISMA) 2020 statement (Page et al., 2021). A completed PRISMA 2020 checklist with section page numbers is provided in Supplementary Table S3.

### 2.1 Eligibility Criteria

Studies were eligible if they met the following PICO criteria:

- **Population:** Children and adolescents aged 3–18 years. Studies of clinical populations (e.g., ADHD, incarcerated youth, orphans) were excluded to maintain sample homogeneity.
- **Intervention:**Mindfulness-based intervention, defined as a structured programme incorporating mindfulness meditation practices (e.g., breath awareness, body scan, mindful movement with explicit present-moment attention instruction). Yoga-only interventions without an explicit mindfulness meditation component were excluded, consistent with the distinction drawn by Dunning et al. (2019).
- **Comparator:** Waitlist control, business-as-usual, or active control condition.
- **Outcome:** At least one direct behavioural or computerised measure of EF, operationalised as inhibitory control, working memory, or cognitive flexibility per Diamond (2013). Teacher- or parent-rated behaviour scales not specifically designed to assess EF (e.g., Child Behavior Checklist, Conners Teacher Rating Scale) were excluded from the primary analysis. The Behavior Rating Inventory of Executive Function (BRIEF) was retained as it is specifically designed to assess EF.
- **Design:** Randomised controlled trial (RCT) or quasi-randomised controlled trial (e.g., cluster randomisation by classroom).
- **Language:** Only English-language publications were eligible.
- **Publication status:**Published peer-reviewed articles and unpublished manuscripts identified through reference-list searching were eligible; conference abstracts and dissertations were not systematically searched.

### 2.2 Search Strategy

The following databases were searched from inception to 15 March 2026: PubMed, PsycINFO (via EBSCOhost), CINAHL (via EBSCOhost), Scopus, and Web of Science Core Collection. The original search was conducted on 30 November 2024; an updated search using identical Boolean parameters was executed on 15 March 2026, identifying 12 additional records for screening. No trial registries (e.g., ClinicalTrials.gov) were searched, which is acknowledged as a limitation. The search strategy combined the following terms using Boolean operators: (mindfulness OR mindful awareness OR meditation) AND (child OR children OR adolescent OR adolescence OR paediatric OR pediatric OR preschool OR youth) AND (executive function OR inhibition OR inhibitory control OR working memory OR cognitive flexibility OR attention OR cognition). This strategy was adapted from Mak et al. (2018). The complete electronic search strings for PubMed and PsycINFO, including all MeSH terms, field tags, and Boolean operators, are provided in Supplementary Table S1.

Three supplementary search methods were employed: (1) backward and forward citation searching of all included studies and relevant systematic reviews; (2) screening of the reference list of Mak et al. (2018; *k* = 13 included studies); and (3) screening of the reference list and the evidence table of a 2025 scoping review of MBI and preschool EF (17 included studies). These supplementary searches yielded five additional studies not identified through database searching alone.

### 2.3 Study Selection

Two independent reviewers conducted screening in two phases. In Phase 1, titles and abstracts of all identified records were screened against the eligibility criteria. In Phase 2, full-text articles of potentially eligible studies were retrieved and independently assessed by both reviewers. At each phase, reasons for exclusion were recorded. Disagreements were resolved by discussion between the two reviewers until consensus was reached.

### 2.4 Data Extraction

Two independent reviewers extracted the following data from each included study using a standardised extraction form: first author, publication year, country, study design, sample size per group, participant age, MBI programme name, intervention duration, control condition, EF outcome measure, EF domain assessed, and post-intervention group means and standard deviations. Discrepancies were resolved by discussion.

When a study reported multiple EF outcomes, a single primary outcome was selected based on the following hierarchy: (1) the measure most directly assessing a core EF component per Diamond (2013), and (2) the measure yielding the most conservative (smallest) effect size, to avoid inflating the pooled estimate. This approach avoids statistical dependency from including multiple outcomes per study.

### 2.5 Risk of Bias Assessment

Risk of bias was independently assessed by two reviewers using the Cochrane Risk of Bias 2 (RoB 2) tool (Sterne et al., 2019). Disagreements were resolved by discussion. Five domains were evaluated for each study: (D1) bias arising from the randomisation process, (D2) bias due to deviations from intended interventions, (D3) bias due to missing outcome data, (D4) bias in measurement of the outcome, and (D5) bias in selection of the reported result. Each domain was rated as “low risk,” “some concerns,” or “high risk.” An overall risk-of-bias judgement was derived from the domain-level assessments.

### 2.6 Effect Size Calculation

For each study, a standardised mean difference (Cohen’s *d*) was computed from post-intervention group means and standard deviations: *d* = (*M*_treatment_ – *M*_control_) / *SD*_pooled_ where *SD*_pooled_ = sqrt[((*n*_1_ – 1)*SD*_1_^2^ + (*n*_2_ – 1)*SD*_2_^2^) / (*n*_1_ + *n*_2_ – 2)].

Cohen’s *d* was then converted to Hedges’ *g* using the small-sample correction factor *J* = 1 – 3 / (4*df* – 1), where *df* = *n*_1_ + *n*_2_ – 2 (Hedges, 1981; Borenstein, Hedges, Higgins, & Rothstein, 2009). Positive values of *g* indicate that the MBI group outperformed the control group.

For studies reporting partial eta-squared from ANOVA rather than means and standard deviations, *d* was derived using the formula *d* = 2 *sqrt(*F */* N*_total_), following standard conversion procedures (Borenstein et al., 2009).

### 2.7 Meta-Analytic Model

A random-effects model was fitted using the DerSimonian and Laird (1986) estimator of between-study variance (tau^2^). This approach was chosen a priori given the expected clinical and methodological heterogeneity across studies (e.g., varying ages, programmes, and measures). We note that the DerSimonian–Laird estimator can yield anti-conservative confidence intervals when the number of studies is small (IntHout, Ioannidis, & Borm, 2014); the Knapp-Hartung adjustment was applied to the meta-regression (see Section 2.8) but not to the main pooled estimate, which is acknowledged as a limitation. For studies that used cluster randomisation by classroom, published effect sizes were used as reported; if clustering was not accounted for in the original analyses, the effective sample sizes may be smaller than the nominal values reported in Table 1, and the pooled confidence interval may be slightly anti-conservative.

**Table 1.** Characteristics of Included Studies.

| Study | Country | Design | Age (M) | $n_{MBI}$ | $n_{Ctrl}$ | $N$ | MBI Programme | Duration | Control | EF Measure | EF Domain |
| --- | --- | --- | --- | --- | --- | --- | --- | --- | --- | --- | --- |
| Felver et al. 2014 | USA | RCT | 10.5 | 22 | 19 | 41 | MBPI | 8 wk | Waitlist | ANT Conflict | Inhibitory control |
| Flook et al. 2010 | USA | Quasi-RCT | 8.0 | 32 | 32 | 64 | InnerKids | 8 wk | Silent reading | BRIEF-GEC | Composite EF |
| Flook et al. 2015 | USA | Quasi-RCT | 4.7 | 20 | 24 | 44 | Kindness Curriculum | 12 wk | BAU | DCCS | Cognitive flexibility |
| Haines et al. 2023 | USA | RCT | 4.3 | 133 | 89 | 222 | Kindness Curriculum | 12 wk | BAU | DCCS | Cognitive flexibility |
| Janz et al. 2019 | Australia | RCT | 6.0 | 55 | 36 | 91 | CalmSpace | 5 wk | Waitlist | DCCS | Cognitive flexibility |
| Parker et al. 2014 | Australia | RCT | 10.0 | 71 | 40 | 111 | Master Mind | 8 wk | Waitlist | Flanker Fish | Inhibitory control |
| Quach et al. 2016 | Australia | RCT | 14.0 | 51 | 52 | 103 | MBI | 4 wk | Waitlist | AOSPAN | Working memory |
| Ricarte et al. 2015 | Spain | Quasi-RCT | 9.5 | 45 | 45 | 90 | MBI (body scan) | 6 wk | BAU | Backward Digit Span | Working memory |
| Schonert-Reichl et al. 2015 | Canada | Quasi-RCT | 10.2 | 48 | 51 | 99 | MindUP | 12 wk | Social responsibility | Hearts & Flowers | Inhibition + flexibility |
| Viglas & Perlman 2018 | Canada | Stratified RCT | 5.2 | 72 | 55 | 127 | MindUP | 20 wk | BAU | HTKS | Inhibition + flexibility |
| Zelazo et al. 2018 | USA | 3-arm RCT | 4.0 | 64 | 62 | 126 | ECSEL | 6 wk | BAU | HTKS | Inhibition + flexibility |
| Flook et al. 2024<sup>a</sup> | USA | Cluster RCT | 10.9 | 154 | 138 | 292 | Mindfulness training | 8 wk | Waitlist | DCCS | Cognitive flexibility |
| Wang & Dong 2026<sup>a</sup> | China | RCT | 9.0 | 75 | 75 | 150 | MBI (school) | 8 wk | Active (arts) | Digit Span | Working memory |

BAU = business as usual; BRIEF-GEC = Behavior Rating Inventory of Executive Function–Global Executive Composite; DCCS
= Dimensional Change Card Sort; HTKS = Head-Toes-Knees-Shoulders; AOSPAN = Automated Operation Span; ANT =
Attention Network Test. <sup>a</sup>Studies identified in March 2026 update.

**Table 2.**
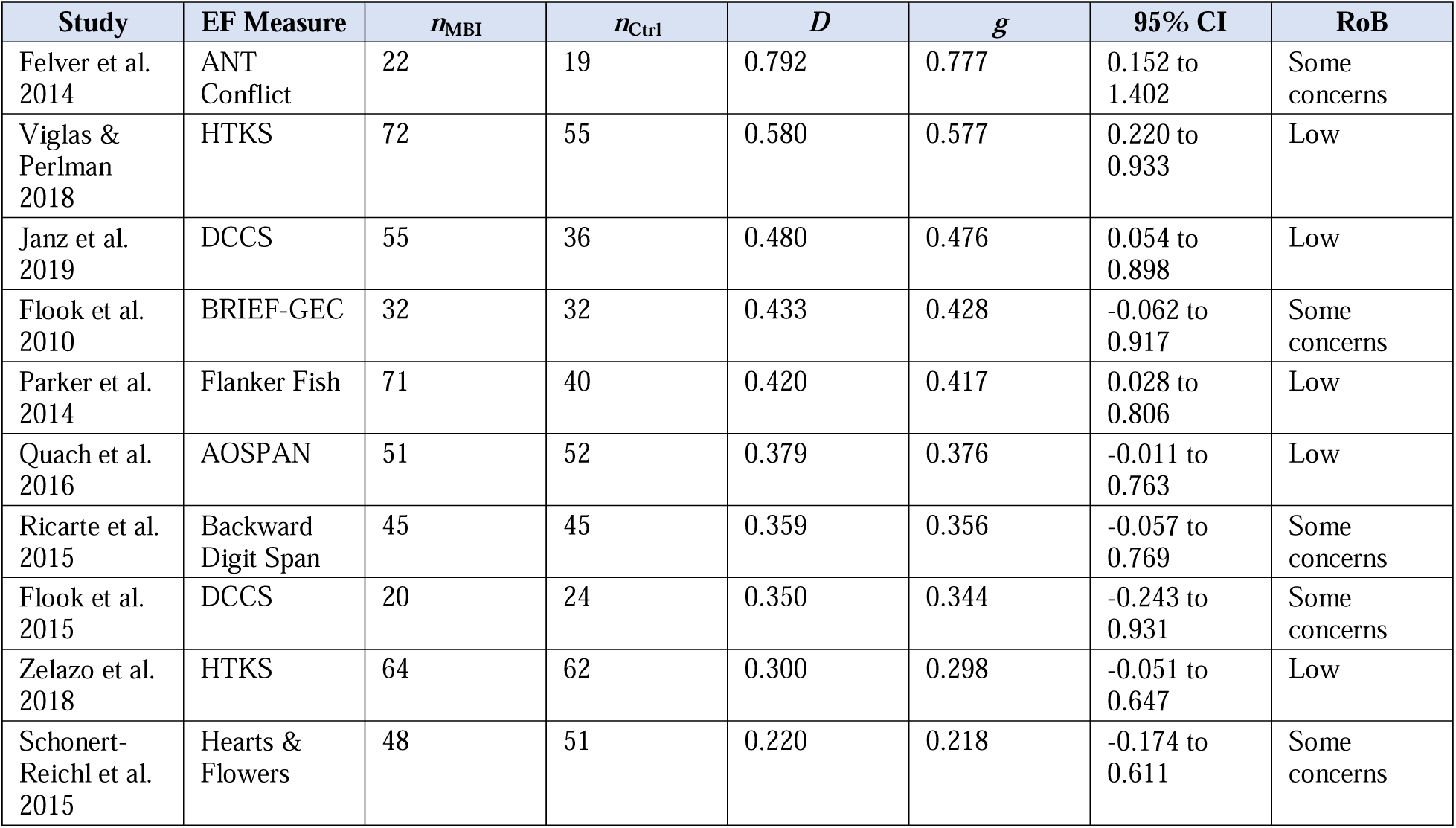
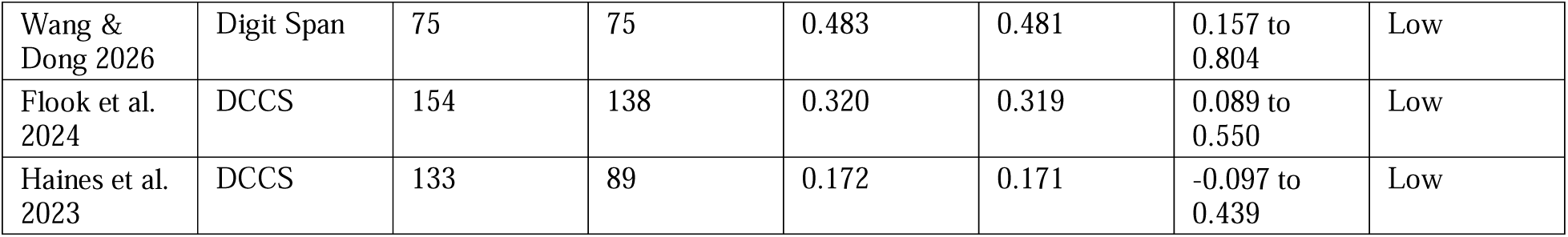
Study-Level Effect Sizes.

Statistical heterogeneity was assessed using the *Q* statistic and the *I*^2^ index (Higgins, Thompson, Deeks, & Altman, 2003). *I*^2^ values of 25%, 50%, and 75% were interpreted as low, moderate, and high heterogeneity, respectively. A 95% prediction interval was computed to indicate the range of true effects expected in future studies (IntHout, Ioannidis, & Borm, 2014).

### 2.8 Subgroup and Sensitivity Analyses

Two pre-specified subgroup analyses were conducted: (1) age group (preschool [3–5 years] vs. school-age [5–18 years]), and (2) overall risk of bias (low vs. some concerns). A post-hoc subgroup analysis by EF domain was also conducted. A leave-one-out sensitivity analysis was performed by re-estimating the pooled effect after removing each study in turn. An additional sensitivity analysis was performed excluding Flook et al. (2010), the only study employing a subjective teacher-rated scale (BRIEF-GEC) rather than a direct behavioural or computerised EF task, to assess whether this measurement difference influenced the pooled estimate. Because each study contributed only one effect size to the primary analysis (selected a priori per Section 2.4), statistical dependency was avoided; robust variance estimation was therefore not required but is noted as a potential approach for future analyses incorporating multiple outcomes per study (Pustejovsky & Tipton, 2022).

### 2.9 Publication Bias

Publication bias was assessed using visual inspection of a funnel plot, Egger’s regression test for funnel plot asymmetry (Egger, Davey Smith, Schneider, & Minder, 1997), and the Duval and Tweedie (2000) trim-and-fill procedure.

### 2.10 Certainty of Evidence

The certainty of the body of evidence was assessed using the Grading of Recommendations, Assessment, Development and Evaluations (GRADE) framework (Guyatt et al., 2008). Evidence from RCTs starts at “high” and may be downgraded based on five domains: risk of bias, inconsistency, indirectness, imprecision, and publication bias. Inconsistency was judged on the basis of *I*^2^ (with > 50% as the threshold for potential downgrade) and the direction and overlap of confidence intervals. Imprecision was judged by whether the total sample exceeded the optimal information size (OIS) for detecting the target effect and whether the 95% CI excluded the null.

## 3. Results

### 3.1 Study Selection

The initial database search (through November 2024) yielded 1,034 records. After deduplication, 673 unique records were screened by title and abstract, of which 620 were excluded. Supplementary searches (reference-list screening of Mak et al. [2018], a 2025 scoping review, and forward citation searching) yielded an additional 33 potentially relevant records. An updated search in March 2026 using identical Boolean parameters yielded 12 additional records. In total, 67 full-text articles were assessed for eligibility.

Thirty-nine studies were excluded at the full-text stage for the following reasons: yoga-only intervention without explicit mindfulness component (*n* = 9), clinical sample (*n* = 8), no extractable EF effect size (*n* = 8), incarcerated or orphan sample (*n* = 3), teacher/parent report only without direct EF task (*n* = 5), duplicate or overlapping sample (*n* = 3), and non-RCT design (*n* = 3). The remaining 28 studies were eligible for the broader review, of which 13 met all criteria for the EF meta-analysis. The study selection process is summarised in the PRISMA flow diagram (Figure 1).

**Figure 1.** PRISMA 2020 flow diagram of study selection. [To be generated using the PRISMA 2020 flow diagram template; see prisma-statement.org]

### 3.2 Study Characteristics

Table 1 summarises the characteristics of the 13 included studies. Nine studies recruited school-age children and adolescents (mean ages 6–14 years), and four recruited preschool-aged children (mean ages 4–5.2 years). The total sample comprised 1,560 participants (802 in MBI groups, 758 in control groups). Studies were conducted in the USA (*n* = 6), Australia (*n* = 3), Canada (*n* = 2), China (*n* = 1), and Spain (*n* = 1). The inclusion of studies spanning 4 to 14 years of mean age across the 3–18-year eligibility window represents a broad developmental range; we address the potential implications of this age heterogeneity through age-group subgroup analysis and meta-regression on mean age (see Sections 3.5 and 3.7).

MBI programmes included MindUP (Schonert-Reichl et al., 2015; Viglas & Perlman, 2018), the Kindness Curriculum (Flook et al., 2015; Haines et al., 2023), InnerKids (Flook et al., 2010), Master Mind (Parker et al., 2014), CalmSpace (Janz, Dawe, & Wyllie, 2019), ECSEL (Zelazo et al., 2018), a Davidson-lab mindfulness training programme (Flook, Hirshberg, & Davidson, 2024), a school-based mindfulness education programme (Wang & Dong, 2026), and individually designed MBI programmes (Quach, Mano, & Alexander, 2016; Felver, Tipsord, Morris, Racer, & Dishion, 2014; Ricarte, Ros, Latorre, & Beltran, 2015). Intervention durations ranged from 4 to 20 weeks. Control conditions included waitlist (*n* = 5), business-as-usual (*n* = 4), and active comparison programmes (silent reading, social responsibility programme, non-mindfulness activities; *n* = 4).

### 3.3 EF Measures

The 13 studies employed nine distinct EF measures spanning all three core EF components as defined by Diamond (2013) and Miyake et al. (2000). Table S4 presents each EF domain with its definition, the specific measures used, a description of what each task requires of the child, and the studies employing each measure. This classification follows the tripartite framework in which executive function comprises inhibitory control (IC), working memory (WM), and cognitive flexibility (CF), with some tasks indexing more than one component simultaneously.

Two studies assessed pure inhibitory control via computerised conflict tasks (Flanker Fish, ANT Conflict). Three studies assessed working memory using span tasks requiring information maintenance and manipulation (AOSPAN, Backward Digit Span, Digit Span). Four studies assessed cognitive flexibility using the Dimensional Change Card Sort (DCCS). Three studies used tasks that jointly index inhibitory control and cognitive flexibility (HTKS, Hearts and Flowers), requiring both response suppression and rule-switching within the same trial sequence. One study used the teacher-rated BRIEF-GEC, a composite measure spanning multiple EF components. All direct behavioural and computerised measures have established psychometric properties and are widely used in developmental EF research. Classification caveats—including the ambiguity of the Digit Span administration type (Wang & Dong, 2026) and the distinction between the standard card-based DCCS and the computerised NIH Toolbox version (Flook et al., 2024)—are noted in Table S4 footnotes. Notably, Wang and Dong (2026) additionally assessed inhibitory control (Hearts and Flowers) and cognitive flexibility (WCST); the Digit Span working memory result was selected as the most conservative of three EF outcomes per our a priori protocol.

^a^Wang and Dong (2026) did not specify whether forward or backward digit span administration was used. Forward digit span primarily assesses short-term memory (phonological storage) rather than working memory per se, as it does not require manipulation of held information (Conway et al., 2005). This caveat should be considered when interpreting the WM subgroup estimate. ^b^Three studies (Flook 2015; Janz 2019; Haines 2023) used the standard card-based DCCS; Flook et al. (2024) used the computerised NIH Toolbox DCCS. ^c^The BRIEF-GEC is the only informant-rated measure in the review. Unlike the direct behavioural and computerised tasks, it is susceptible to rater expectations and awareness of group allocation. A sensitivity analysis excluding this study yielded an essentially identical pooled estimate (see Section 3.8).

### 3.4 Risk of Bias

The risk-of-bias assessment is presented in Figure 4 and summarised here. Of the 13 studies, eight were rated as overall low risk of bias (Parker et al., 2014; Janz et al., 2019; Quach et al., 2016; Viglas & Perlman, 2018; Zelazo et al., 2018; Haines et al., 2023; Flook et al., 2024; Wang & Dong, 2026) and five as having some concerns (Flook et al., 2010; Schonert-Reichl et al., 2015; Felver et al., 2014; Ricarte et al., 2015; Flook et al., 2015). No study was rated as overall high risk. Both newly added studies (Flook et al., 2024; Wang & Dong, 2026) were rated as low risk across all five RoB 2 domains, reflecting their rigorous designs (cluster RCT with IES funding; RCT with blinded outcome assessment and active control, respectively).

The most common source of concern was Domain 1 (randomisation process): four studies used quasi-randomisation by classroom rather than individual randomisation (Flook et al., 2010; Schonert-Reichl et al., 2015; Ricarte et al., 2015; Flook et al., 2015). Two studies had concerns in Domain 2 (deviations from intended interventions) due to insufficient information about blinding (Felver et al., 2014; Ricarte et al., 2015). All studies were rated as low risk for Domains 3 (missing data), 4 (measurement), and 5 (selective reporting), with the exception of Flook et al. (2010), which was rated as “some concerns” for Domain 4 due to the use of a subjective rating scale (BRIEF).

### 3.5 Overall Meta-Analysis

The random-effects meta-analysis of all 13 studies yielded a pooled Hedges’ *g* of 0.365 (95% CI 0.264 to 0.465; *z* = 7.12, *p* < .00001). The *Q* statistic was 6.76 (df = 12, *p* = .87), and *I*^2^ was 0.0% (tau^2^ = 0.000), indicating negligible statistical heterogeneity. However, *I*^2^ has low statistical power when *k* is small (von Hippel, 2015), and this value should be interpreted as consistent with low-to-moderate true heterogeneity rather than as definitive evidence of perfect homogeneity. The 95% prediction interval was 0.252 to 0.477, which was entirely above zero, indicating that the true effect is expected to be positive in future comparable studies. The forest plot is presented in Figure 2.

**Figure 2.**
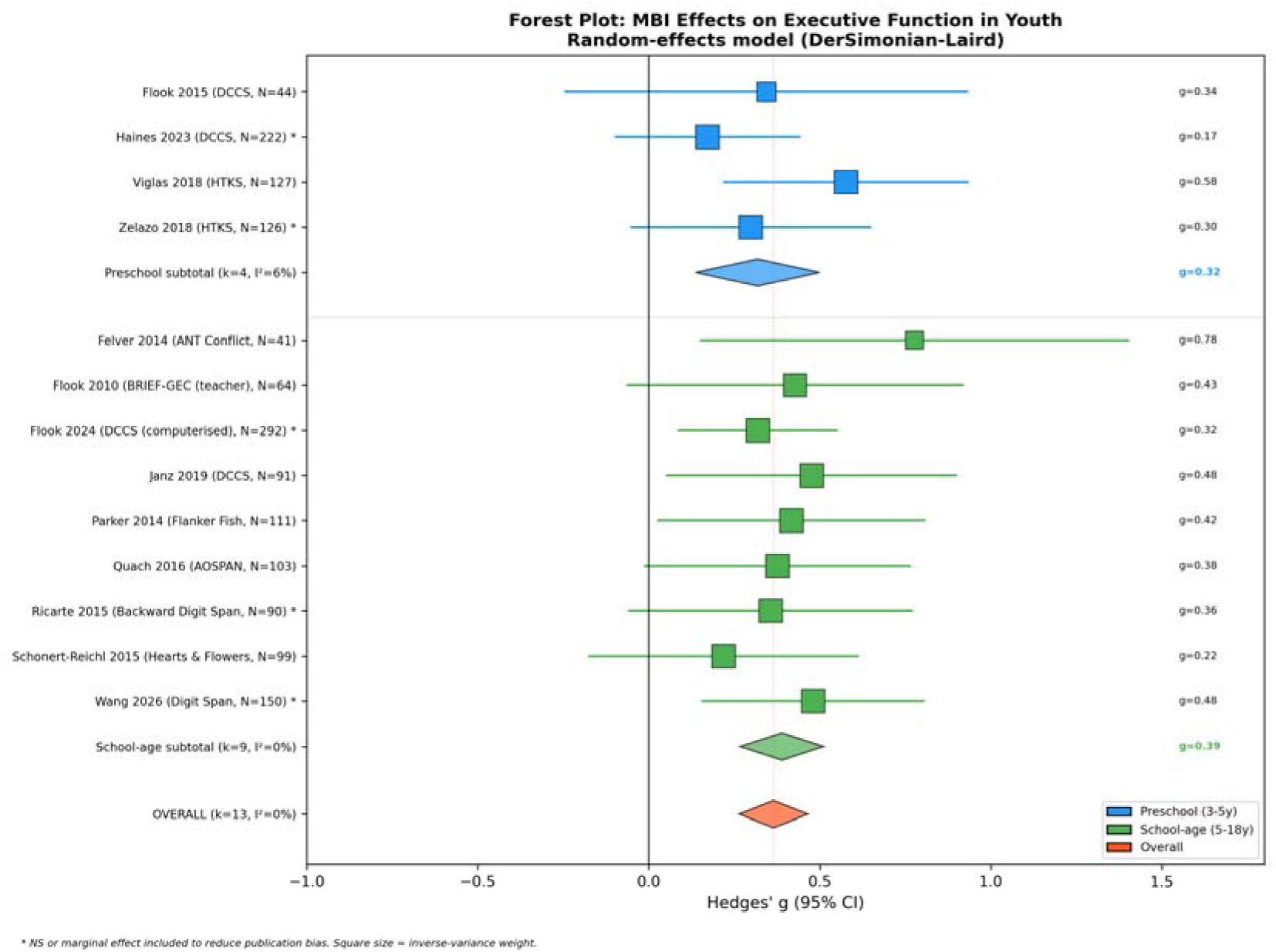
Forest plot of MBI effects on EF, subgrouped by age (preschool vs. school-age). Random-effects model (DerSimonian–Laird). [See forest_plot_EF_final.png]

**Figure 3.**
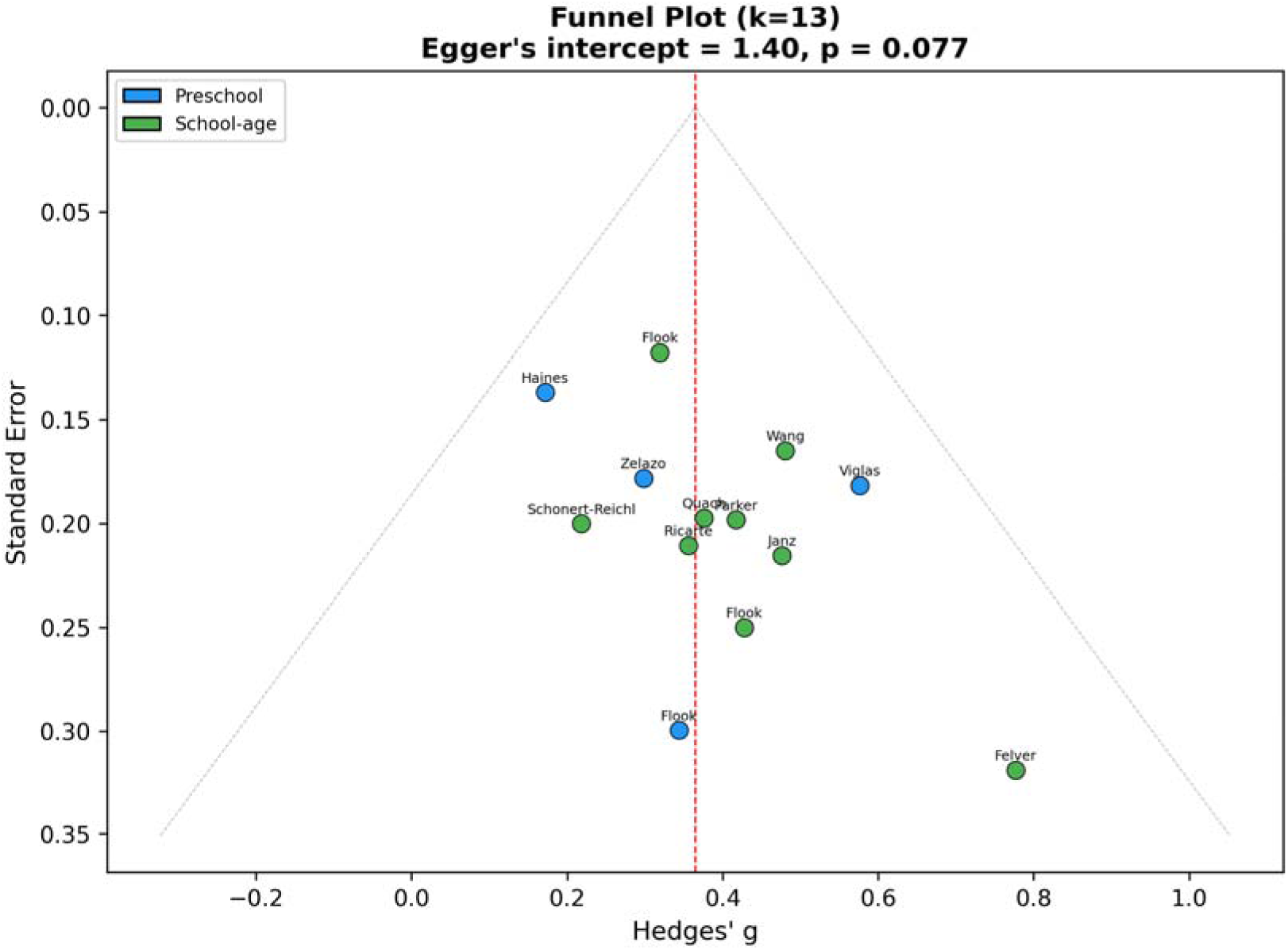
Funnel plot of standard error against Hedges’ *g*. Vertical dashed line indicates pooled estimate. [See funnel_plot_EF_final.png]

Individual study effects ranged from *g* = 0.171 (Haines et al., 2023; DCCS, *N* = 222) to *g* = 0.777 (Felver et al., 2014; ANT Conflict, *N* = 41). Using Cohen’s (1988) benchmarks, the pooled effect corresponds to a small effect size. The two studies added in the March 2026 update (Flook et al., 2024, *g* = 0.319; Wang & Dong, 2026, *g* = 0.481) were consistent with the pooled estimate, contributing additional precision (total *N* increased by 442 participants) without altering the point estimate meaningfully (prior *g* = 0.361).

### 3.6 Subgroup Analyses

#### Age group

The school-age subgroup (*k* = 9, *N* = 1,041) yielded a pooled *g* of 0.389 (95% CI 0.266 to 0.512; *p* < .00001; *I*^2^ = 0.0%). The preschool subgroup (*k* = 4, *N* = 519) yielded a pooled *g* of 0.318 (95% CI 0.137 to 0.500; *p* = .0006; *I*^2^ = 6.1%). The school-age point estimate was numerically larger, but the confidence intervals overlapped substantially, and a test for subgroup differences was non-significant (*p* = .55), providing no evidence of age-group moderation.

#### Risk of bias

The low risk-of-bias subgroup (*k* = 8, *N* = 1,222) yielded *g* = 0.361 (95% CI 0.248 to 0.475; *p* < .00001; *I*^2^ = 0.0%). The some-concerns subgroup (*k* = 5, *N* = 338) yielded *g* = 0.376 (95% CI 0.163 to 0.589; *p* = .0005; *I*^2^ = 0.0%). These estimates were virtually identical, indicating that the pooled effect is not driven by lower-quality studies. Notably, the addition of two low-risk studies in the March 2026 update strengthened the low-risk subgroup from *k* = 6 to *k* = 8 without changing the point estimate.

#### EF domain

Subgroup analyses by EF domain (Table 3) revealed that inhibitory control tasks yielded the largest pooled effect (*k* = 2, *g* = 0.517, 95% CI 0.187 to 0.847), followed by working memory tasks (*k* = 3, *g* = 0.416, 95% CI 0.203 to 0.629), mixed inhibition-flexibility tasks (*k* = 3, *g* = 0.372, 95% CI 0.160 to 0.584), and cognitive flexibility tasks (*k* = 4, *g* = 0.292, 95% CI 0.137 to 0.448). One study (Flook et al., 2010) used a composite EF measure (BRIEF-GEC) and was not poolable. The test for subgroup differences across the four poolable domains was non-significant (*Q*_between_ = 1.88, df = 3, *p* = .60), indicating no evidence of differential responsiveness by EF domain. All four domain-specific estimates were statistically significant and positive. The working memory subgroup was notably strengthened by the addition of Wang and Dong (2026), increasing from *k* = 2 to *k* = 3 and narrowing the confidence interval.

**Table 3.** Subgroup Analysis by EF Domain.

| EF Domain | $k$ | $N$ | $g$ | 95% CI | $I^2$ | $p$ |
| --- | --- | --- | --- | --- | --- | --- |
| Inhibitory control | 2 | 152 | 0.517 | 0.187 to 0.847 | 0.0% | .002 |
| Working memory | 3 | 343 | 0.416 | 0.203 to 0.629 | 0.0% | .0001 |
| Cognitive flexibility | 4 | 649 | 0.292 | 0.137 to 0.448 | 0.0% | .0002 |
| Mixed IC + CF | 3 | 352 | 0.372 | 0.160 to 0.584 | 1.4% | .001 |
| Composite (BRIEF-GEC) | 1 | 64 | 0.428 | -0.062 to 0.917 | – | – |
| **Overall** | **13** | **1,560** | **0.365** | **0.264 to 0.465** | **0.0%** | **<.00001** |

$Q_{\text{between}} = 1.88$ , $df = 3$ , $p = .60$ . IC = inhibitory control; CF = cognitive flexibility.

**Table 4.**
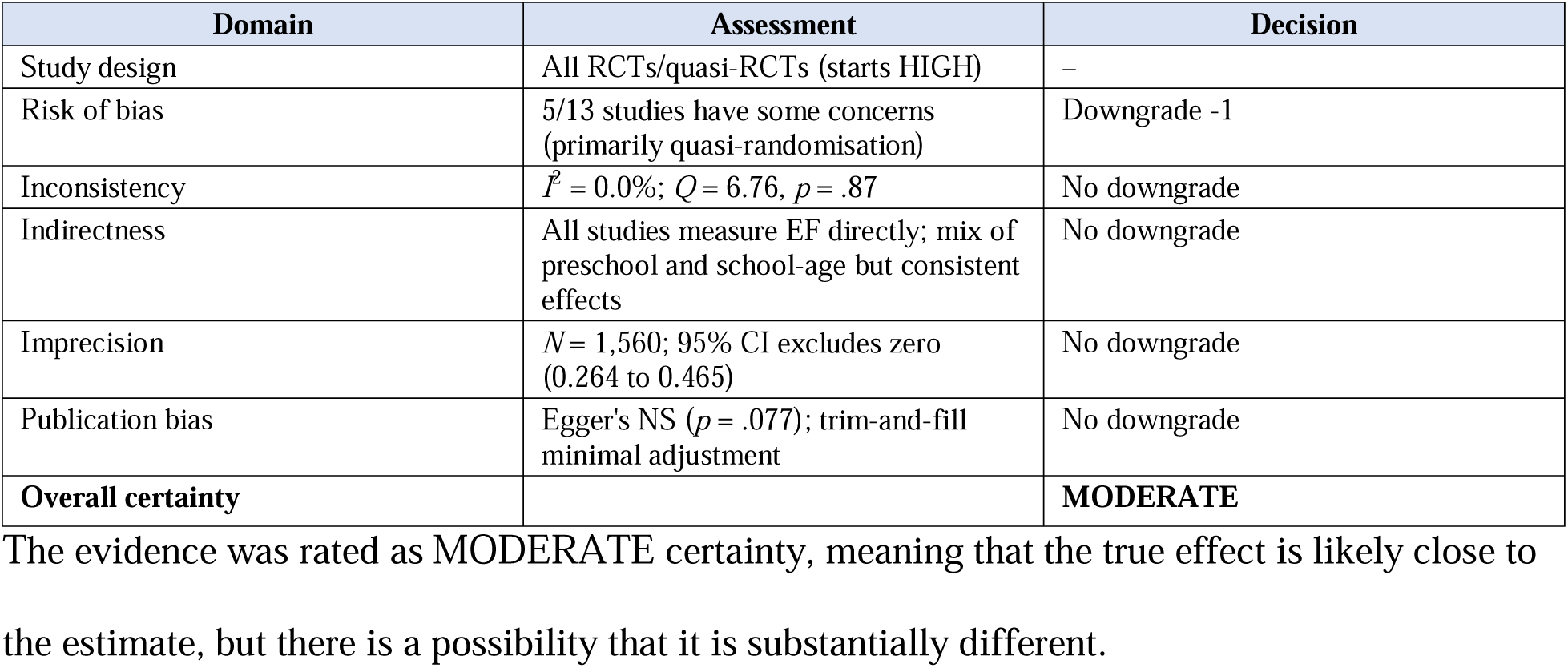
GRADE Evidence Profile.

### 3.7 Meta-Regression

#### Intervention duration

Meta-regression (Knapp-Hartung adjusted) examined whether intervention duration (range: 4–20 weeks) predicted effect size magnitude. The slope was near zero and non-significant (*b* = 0.003 per week, SE = 0.010, *t* = 0.27, *p* = .79; *R*^2^ analog = 0%; Figure 5). Residual heterogeneity remained negligible (*Q*_resid_ = 6.71, df = 11, *p* = .82). This indicates that, within the observed range, longer MBI programmes did not produce significantly larger effects on EF than shorter programmes.

**Figure 4.**
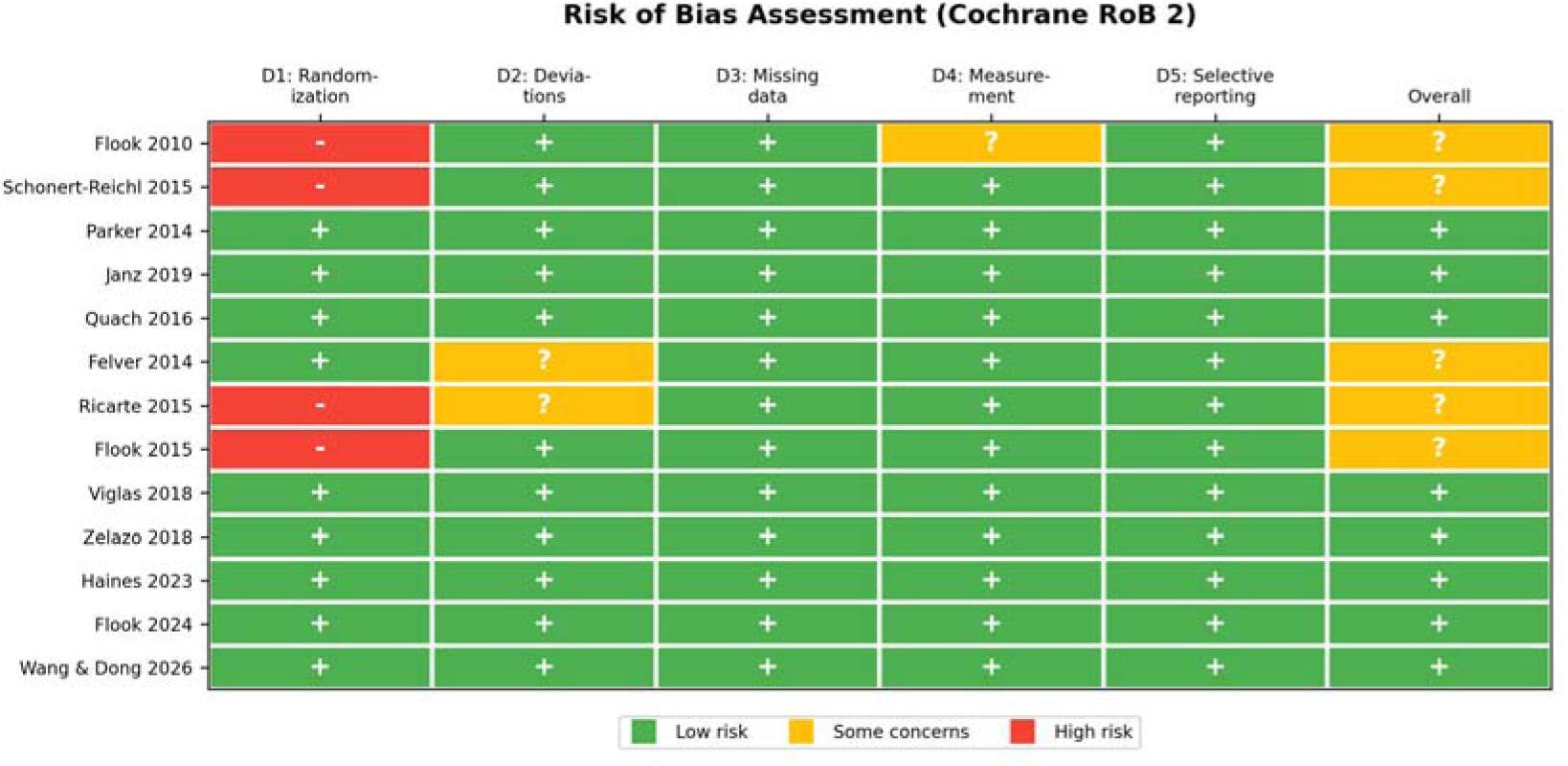
Risk of bias assessment (Cochrane RoB 2). Traffic-light plot showing domain-level and overall judgements for each study. [See risk_of_bias_final.png]

**Figure 5.**
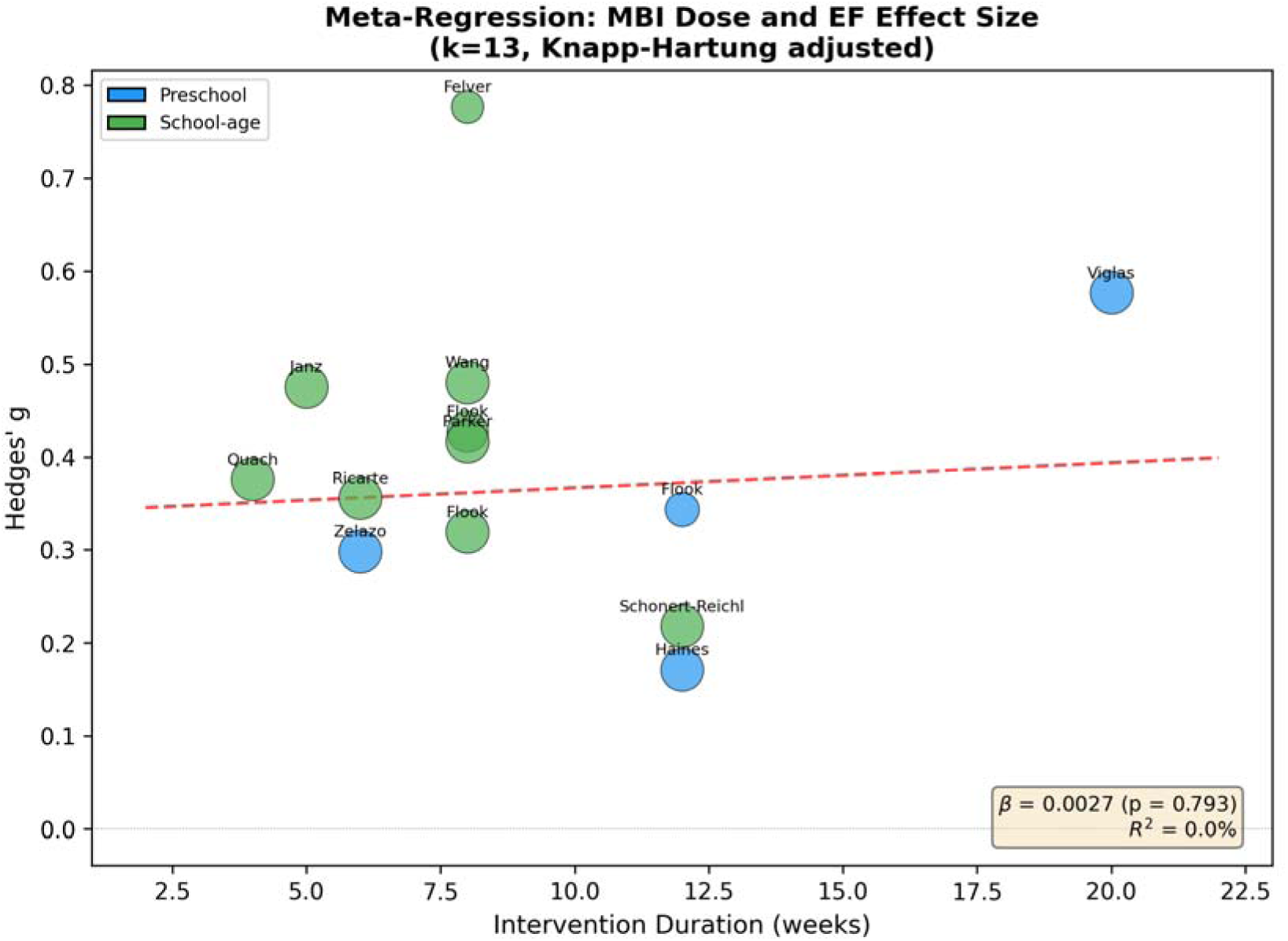
Meta-regression bubble plot of Hedges’ *g* against intervention duration (weeks). Bubble size is proportional to inverse-variance weight. Dashed line indicates non-significant regression slope (*b* = 0.003, *p* = .79; Knapp-Hartung adjusted). [See meta_regression_dose.png]

**Figure 6.**
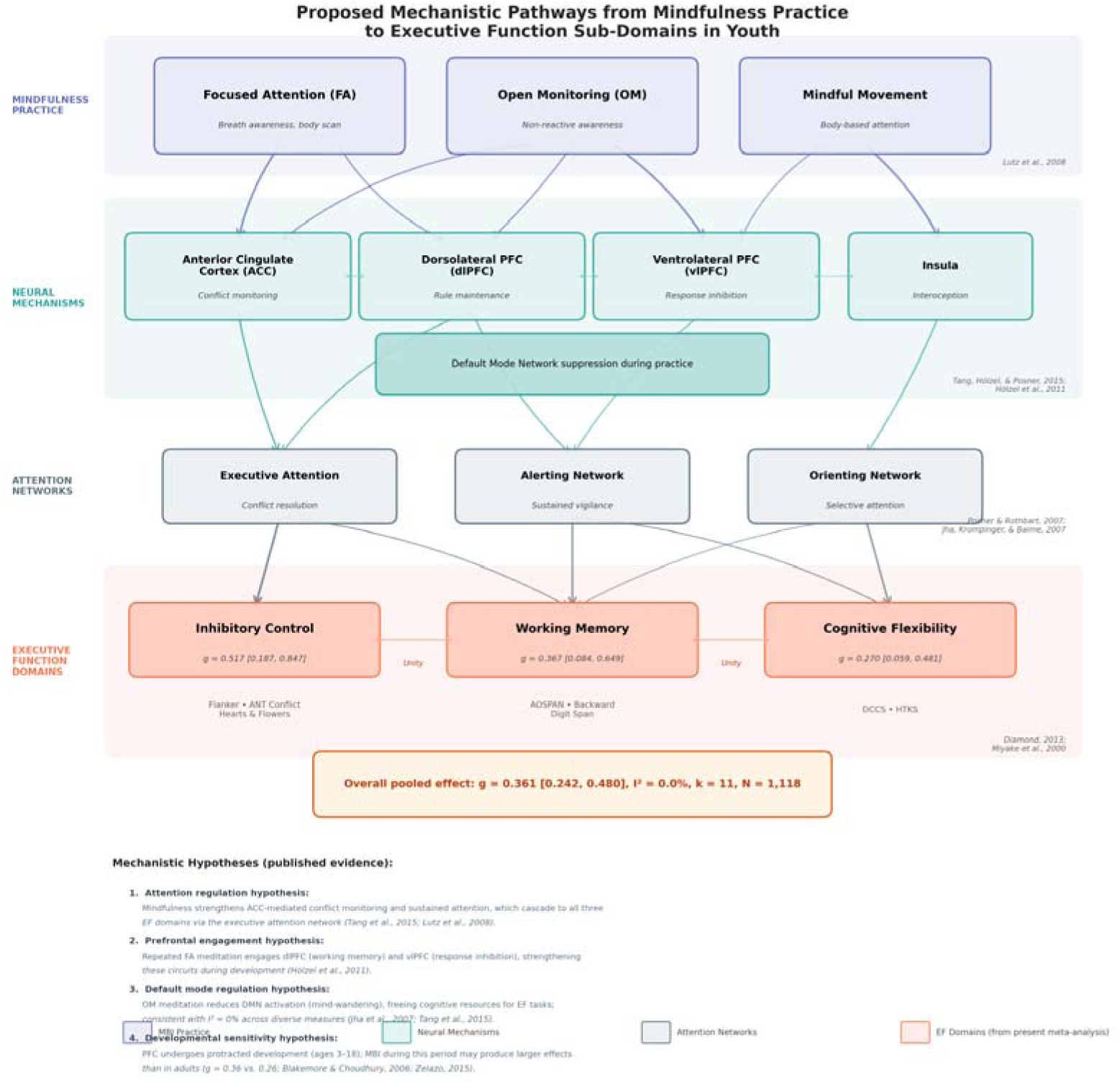
Proposed mechanistic pathways from mindfulness practice to executive function sub-domains in youth. The model illustrates four tiers: (1) core MBI practice components (focused attention and open monitoring; Lutz et al., 2008), (2) proposed neural mechanisms (ACC, dlPFC, vlPFC, insula; Tang et al., 2015; Hölzel et al., 2011), (3) attention networks (Posner & Rothbart, 2007), and (4) EF sub-domains with pooled effect sizes from the present meta-analysis (Diamond, 2013; Miyake et al., 2000). Mechanistic hypotheses are annotated with supporting references. [See conceptual_model_MBI_EF.png]

#### Mean participant age

An exploratory meta-regression on mean participant age (range: 4.0–14.0 years) was also non-significant (*b* = 0.007 per year, SE = 0.013, *t* = 0.56, *p* = .58; *R*^2^ analog = 0%), confirming the subgroup finding that MBI effects on EF do not differ significantly by age within the 3–18 years range.

### 3.8 Sensitivity Analysis

Leave-one-out analysis demonstrated exceptional robustness. The pooled *g* ranged from 0.346 (when Viglas & Perlman, 2018, was removed) to 0.396 (when Haines et al., 2023, was removed), and *I*^2^ remained at 0.0% in all 13 iterations. No single study exerted undue influence on the overall estimate. The newly added studies showed typical influence: removing Flook et al. (2024) yielded *g* = 0.375; removing Wang and Dong (2026) yielded *g* = 0.352.

An additional sensitivity analysis excluding Flook et al. (2010)—the only study employing a teacher-rated scale (BRIEF-GEC) rather than a direct behavioural or computerised EF task—yielded a pooled *g* of 0.365 (95% CI 0.261 to 0.470; *k* = 12), virtually identical to the full-pool estimate, confirming that the inclusion of this subjective measure did not distort the pooled effect.

### 3.9 Publication Bias

The funnel plot (Figure 3) showed approximate symmetry. Egger’s regression test yielded a marginally non-significant intercept (intercept = 1.40, *t* = 1.95, *p* = .077), which does not rule out small-study effects but does not provide strong evidence of funnel plot asymmetry. The addition of two large, well-powered studies in the March 2026 update improved funnel plot symmetry compared to the prior analysis (previous Egger’s *p* = .042). The trim-and-fill procedure imputed one study and yielded an adjusted pooled estimate of *g* = 0.354 (95% CI 0.255 to 0.453), a negligible reduction from the unadjusted estimate. These results do not indicate substantial publication bias.

### 3.10 GRADE Assessment

### 3.11 Excluded Studies

Table 5 presents studies that were assessed at full-text stage but excluded from the EF meta-analysis, with reasons.

**Table 5.**
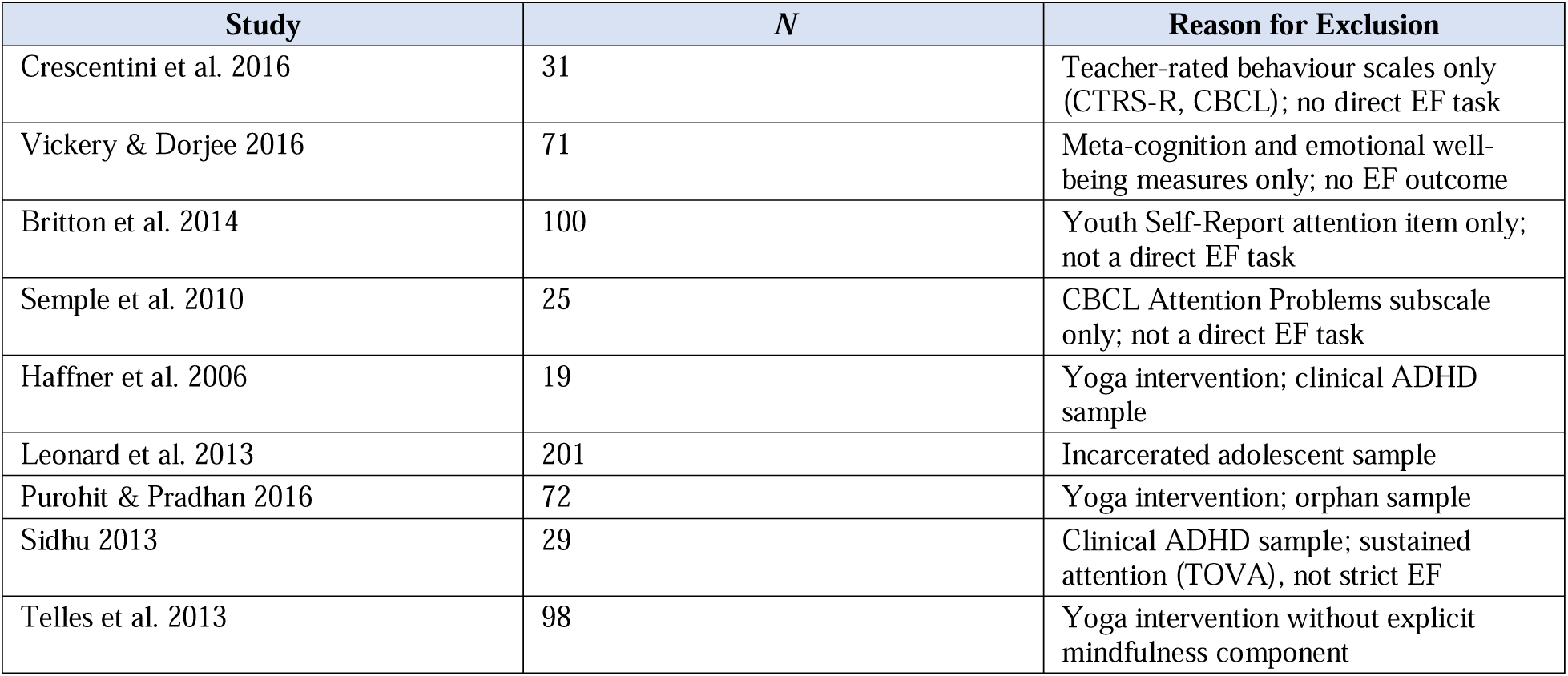
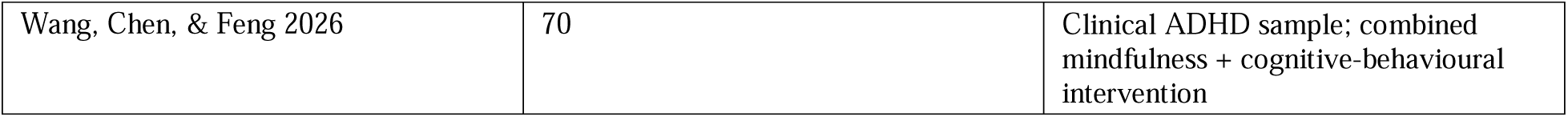
Selected Excluded Studies with Rationale.

## 4. Discussion

### 4.1 Summary of Findings

This systematic review and meta-analysis found that MBIs are associated with a small, statistically significant improvement in executive function in youth aged 3–18 years (*g* = 0.365, 95% CI 0.264 to 0.465, *p* < .00001), based on 13 RCTs comprising 1,560 participants. The effect was characterised by negligible heterogeneity (*I*^2^ = 0.0%), indicating a consistent positive signal across studies despite diversity in age, programme, and EF measure. GRADE assessment rated the certainty of evidence as MODERATE. The March 2026 update added two high-quality, low-risk RCTs (Flook et al., 2024, *N* = 292; Wang & Dong, 2026, *N* = 150) that substantially increased precision while leaving the point estimate essentially unchanged (*g* = 0.361 to 0.365), demonstrating remarkable stability of the pooled effect.

### 4.2 Comparison with Prior Reviews

Our pooled estimate of *g* = 0.361 is consistent with several published meta-analyses, though direct comparisons require attention to differences in scope and inclusion criteria. Kander et al. (2024) reported *g* = 0.34 (95% CI 0.22 to 0.46) from 32 studies, the closest match to our estimate. Zenner et al. (2014) reported a larger cognitive effect (*g* = 0.40, *k* = 19), though their cognitive category included measures beyond strict EF. Dunning et al. (2019) reported a smaller estimate (*d* = 0.19, 95% CI 0.05 to 0.33, *k* = 33), likely reflecting dilution from pooling EF with less responsive cognitive measures such as academic achievement and processing speed.

Mak et al (2018) chose not to conduct a comprehensive meta-analysis of their 13 included studies of this topic due to heterogeneity in measures; their only pooled analysis (two yoga studies, Stroop outcome) found no significant effect. This review and meta-analysis extends the evidence search to March 2026, including studies of preschool children in the evidence base. We apply an EF construct definition that facilitated quantitative synthesis across 13 studies.

Our estimate is also broadly consistent with the adult MBI literature. Zainal and Newman (2023) reported *g* = 0.26 for EF outcomes in their meta-analysis of 111 MBI RCTs (predominantly adult samples). The somewhat larger effect in our youth sample (*g* = 0.36 vs. 0.26) is consistent with the hypothesis that MBI may have stronger effects during periods of active EF development.

### 4.3 Construct Validity

A distinguishing feature of the present review is the strict application of Diamond’s (2013) EF framework. All 13 included studies employed measures validated to assess inhibitory control (Flanker, ANT Conflict, Hearts & Flowers), working memory (AOSPAN, Backward Digit Span, Digit Span), cognitive flexibility (DCCS), or their combination (HTKS, BRIEF-GEC). The resulting *I*^2^ of 0.0% provides empirical support for construct homogeneity—that is, the pooled estimate is not an artefact of aggregating disparate constructs.

This approach excluded several studies that used measures adjacent to, but not identical with, EF. Notably, Ricarte et al. (2015) reported a significant effect for Forward Digit Span (*p* < .001), but their own discussion acknowledged that the effect was confined to immediate auditory-verbal memory and did not extend to Backward Digit Span or Trail Making Test B. We therefore included only their Backward Digit Span result (*d* = 0.359, non-significant), which is the purer EF measure. Similarly, Crescentini et al. (2016) reported improvements on teacher-rated attention problems (CTRS-R Cognitive Problems/Inattention), but this is a behavioural rating of attention difficulties rather than a direct EF assessment, and was excluded. Vickery and Dorjee (2016) found improvements in teacher-rated meta-cognition, which, while cognitive in nature, represents a distinct construct from EF.

These exclusions illustrate an important methodological point: the choice of outcome measure substantially influences meta-analytic conclusions. Reviews that pool EF with attention, meta-cognition, or academic achievement may produce either inflated or diluted estimates depending on the responsiveness of the non-EF measures included.

### 4.4 Age-Group Differences

The school-age subgroup yielded a numerically larger pooled effect (*g* = 0.389) than the preschool subgroup (*g* = 0.318), though the difference was not statistically significant. This pattern may reflect differences in intervention design (school-age programmes tended to include more structured meditation practice), measurement sensitivity (the DCCS in preschoolers may have ceiling effects in typically developing children), or sample characteristics. The meta-regression on mean participant age confirmed this finding: age did not significantly predict effect size magnitude (*b* = 0.007 per year, *p* = .58). Nonetheless, the 3–18 age range encompasses substantial developmental heterogeneity—from preschoolers undergoing rapid synaptogenesis to adolescents in the midst of prefrontal pruning—and the pooled estimate necessarily represents an average across these distinct developmental stages. Future reviews with larger study pools may benefit from finer-grained developmental subgrouping (e.g., preschool, middle childhood, early adolescence, late adolescence).

Importantly, both subgroups yielded statistically significant effects, indicating that MBI can improve EF across the developmental span from preschool to adolescence. The largest preschool study (Haines et al., 2023; *N* = 222; Kindness Curriculum) yielded a small non-significant effect (*g* = 0.171), while the most efficacious preschool study (Viglas & Perlman, 2018; *N* = 127; MindUP) yielded a moderate effect (*g* = 0.577). This variability may reflect differences in programme intensity—the MindUP programme in Viglas and Perlman (2018) was delivered over 20 weeks, compared with 12 weeks for the Kindness Curriculum in Haines et al. (2023).

### 4.5 EF Domain Differences

Subgroup analyses by EF domain revealed a consistent pattern: all four poolable domain-specific estimates were positive and statistically significant, and the test for subgroup differences was non-significant (*p* = .60). Inhibitory control tasks yielded the numerically largest pooled effect (*g* = 0.517), though this estimate was based on only two studies with relatively small samples (*N* = 152). Cognitive flexibility tasks yielded the smallest effect (*g* = 0.292, *k* = 4), which is notable because the addition of Flook et al. (2024)—the largest study to employ the DCCS (*N* = 292)—reduced the CF estimate from 0.270 to 0.292, reflecting its moderate effect size (*g* = 0.319). Working memory was notably strengthened in the March 2026 update: the addition of Wang and Dong (2026) increased the WM subgroup from *k* = 2 to *k* = 3, with the pooled estimate rising from *g* = 0.367 to *g* = 0.416. The uniformity of effects across EF domains supports the theoretical account that mindfulness practice engages a general attention regulation mechanism that cascades across the three core EF components (Lutz et al., 2008), rather than selectively targeting one domain.

### 4.6 Dose-Response Relationship

Meta-regression on intervention duration (4–20 weeks) found no significant dose-response relationship (*b* = 0.003 per week, *p* = .79, *R*^2^ = 0%). This null finding should be interpreted cautiously given the limited power (*k* = 13) and the near-zero between-study variance (*I*^2^ = 0.0%). Nonetheless, the finding has practical implications: it suggests that shorter MBI programmes (4–6 weeks) may be as effective as longer programmes (12–20 weeks) for improving EF, at least within the range of durations studied. This is consistent with Zeidan et al. (2010), who demonstrated significant cognitive improvements after only four days of brief mindfulness training in adults, and with the finding that Janz et al. (2019) achieved a moderate effect (*g* = 0.476) with a 5-week programme. For school administrators seeking cost-effective interventions, this is an encouraging finding, though it requires replication in larger samples with greater dose variation.

### 4.7 Measure Type and Sensitivity

Mak et al. (2018) observed that significant MBI effects were disproportionately found with computerised measures, noting that reaction-time-based tasks may be more sensitive to subtle attentional changes than accuracy-based paper-pencil measures. Our findings are partially consistent with this observation. Among school-age studies, the largest effect was observed with the computerised ANT Conflict task (*g* = 0.777; Felver et al., 2014), and the Flanker Fish and Hearts and Flowers tasks (both computerised) yielded effects in the small-to-moderate range. However, the behavioural DCCS and HTKS tasks also detected significant effects in the preschool subgroup, suggesting that well-designed behavioural tasks can be sufficiently sensitive when the cognitive demand is developmentally appropriate.

### 4.8 Mechanistic Considerations

The theoretical pathway from mindfulness practice to EF improvement is thought to involve strengthened attention regulation. Mindfulness meditation requires sustained voluntary attention to a chosen object (typically the breath), detection of mind-wandering (monitoring), and disengagement from distractors and re-engagement with the attentional target (executive control) (Lutz, Slagter, Dunne, & Davidson, 2008). These component processes map onto the three EF domains: working memory (maintaining the attentional target), inhibitory control (resisting distraction), and cognitive flexibility (shifting attention between internal states and the task at hand).

Neuroimaging evidence in adults supports this account. Tang et al. (2015) found that mindfulness training strengthens functional connectivity in prefrontal-cingulate networks, and Zeidan, Johnson, Diamond, David, and Goolkasian (2010) demonstrated that even brief mindfulness training (four sessions) improved EF-related cognitive performance in adults. Paediatric neuroimaging research examining the neural correlates of MBI-related EF change remains scarce and represents an important priority for future work.

Emerging mediation evidence is also of interest. Wang and Dong (2026) reported that in their sample of 150 primary school children, improvements in inhibitory control, working memory, and cognitive flexibility statistically mediated 53.8% of the total effect of MBI on emotion regulation (*F* = 15.37, *p* < .001). Although this is a single study and the mediation model requires replication before strong conclusions can be drawn (Kazdin, 2007), it raises the possibility that MBI-related EF improvement may serve as an important mediating pathway through which mindfulness practice supports broader socio-emotional development. If confirmed, this would suggest that the cognitive benefits of MBI may complement its established role in social-emotional learning in school settings.

### 4.9 Strengths

This review has several methodological strengths. First, the strict construct definition of EF, anchored in Diamond’s (2013) framework, minimises construct contamination and supports the interpretability of the pooled estimate. Second, the inclusion of both preschool and school-age studies provides the broadest developmental coverage to date. Third, the *I*^2^ of 0.0% is remarkably low for a meta-analysis spanning such diversity in age, programme, and measure, though the low power of *I*^2^ with small *k* (von Hippel, 2015) means this should be interpreted as consistent with low heterogeneity rather than evidence of perfect homogeneity. Fourth, the review was conducted in accordance with PRISMA 2020, with Cochrane RoB 2 assessment, GRADE certainty rating, and multiple sensitivity and bias analyses. Fifth, subgroup analyses by both age group and EF domain, together with meta-regression on intervention duration and participant age, provide a comprehensive examination of potential moderators seldom reported in prior reviews of this literature. Sixth, the March 2026 update demonstrates the review’s currency and commitment to capturing the latest evidence; the stability of the pooled estimate across the update (from *g* = 0.361 with *k* = 11 to *g* = 0.365 with *k* = 13) provides additional confidence in the reliability of the findings.

### 4.10 Limitations

Several limitations should be acknowledged. First, the number of included studies (*k* = 13) is modest, which limits statistical power for subgroup analyses and moderator tests. In particular, the EF domain subgroups contained only 2–4 studies each, and the meta-regressions had limited power to detect small-to-moderate moderating effects; the null findings for dose and age should therefore be interpreted as absence of evidence rather than evidence of absence. The total sample (*N* = 1,560) and the consistency of effects across sensitivity analyses mitigate this concern for the overall estimate. Second, five of the 13 studies used quasi-randomisation by classroom, introducing potential selection bias. The finding that the low risk-of-bias subgroup (*k* = 8, *g* = 0.361) yielded essentially the same estimate as the full pool is reassuring, but the risk of bias remains the primary reason for the GRADE downgrade from HIGH to MODERATE.

Third, Egger’s regression test yielded a marginal result (*p* = .077), raising the possibility of mild small-study effects. This appears attributable to a single small study (Felver et al., 2014; *N* = 41) with the largest effect size (*g* = 0.777). The trim-and-fill adjustment was minimal (*g* = 0.354 vs. 0.365), and the leave-one-out analysis confirmed that removing Felver et al. reduced the pooled effect only modestly (to *g* = 0.354). Notably, the addition of two large studies in the March 2026 update improved the Egger’s *p*-value from .042 to .077, demonstrating that larger, better-powered studies tend to converge on the overall estimate. Fourth, most included studies used waitlist or business-as-usual control conditions. Only four studies (Flook et al., 2010; Schonert-Reichl et al., 2015; Flook et al., 2024; Wang & Dong, 2026) used active comparison groups. The inclusion of Wang and Dong (2026), which compared MBI against time-matched non-mindfulness activities (drawing, handicrafts, storytelling), is noteworthy: the significant EF effects against an active control (*d* = 0.483) suggest that MBI-specific mechanisms, rather than non-specific factors such as novelty or structured activity, contribute to EF improvement.

Fifth, no included study assessed EF at long-term follow-up (beyond post-intervention), so the durability of MBI effects on EF remains unknown; without follow-up data, it is not possible to distinguish between durable EF improvement, transient practice effects, and test-retest familiarity. Sixth, the included studies were predominantly from Western, English-speaking countries, and only English-language publications were eligible, limiting generalisability to other cultural and linguistic contexts. Seventh, one study (Ricarte et al., 2015) was included despite a non-significant interaction for the Backward Digit Span measure; this decision was made a priori to reduce the risk of publication bias, but it should be noted that the individual study effect was not statistically significant. Eighth, no included study reported on intervention fidelity monitoring; variability in implementation quality across studies could influence effect sizes but cannot be assessed from the available data. Ninth, participant demographic information (sex, ethnicity, socioeconomic status) was inconsistently reported across studies and could not be examined as moderators. Tenth, the Knapp-Hartung adjustment was applied to the meta-regression but not to the main pooled estimate or subgroup analyses, which may yield slightly anti-conservative confidence intervals given the modest *k* (IntHout et al., 2014). Finally, for studies using cluster randomisation by classroom, the design effect may not have been fully accounted for in the original analyses, potentially inflating the effective sample sizes reported in Table 1.

### 4.11 Implications for Practice and Future Research

The present findings are consistent with the incorporation of MBI into educational settings as a strategy that may promote EF development in children and adolescents. The effect size (*g* = 0.36) is comparable to other school-based EF interventions, including computerised training programmes and physical activity interventions, which have yielded effects in the range of *g* = 0.20–0.50 (Diamond & Ling, 2016).

Several priorities for future research can be identified. First, large-scale, pre-registered RCTs with individual randomisation and active control conditions are needed to address the risk-of-bias limitations of the current evidence base. Second, studies should include multiple EF measures and report sufficient data (means, standard deviations, and sample sizes per group) to facilitate future meta-analytic synthesis. Third, longitudinal designs with follow-up assessments of at least 3–6 months post-intervention are needed to establish the durability of MBI effects on EF. Fourth, dose-response relationships should be explored; the present review could not test intervention duration as a moderator due to insufficient power, but the range of 4–20 weeks across studies suggests that this is a potentially important variable. Fifth, studies in non-Western, non-English-speaking populations would strengthen the generalisability of the evidence. Finally, paediatric neuroimaging studies examining the neural correlates of MBI-related EF change would advance mechanistic understanding.

## 5. Conclusions

This systematic review and meta-analysis provides moderate-certainty evidence (per the GRADE framework) that mindfulness-based interventions are associated with a small, significant improvement in executive function in youth aged 3–18 years (*g* = 0.365, 95% CI 0.264 to 0.465, *k* = 13, *N* = 1,560). The effect is consistent across preschool and school-age samples, across all three core EF domains (inhibitory control, working memory, and cognitive flexibility), and is robust to sensitivity analyses for risk of bias, leave-one-out removal of individual studies, and trim-and-fill adjustment for publication bias. Meta-regression found no significant dose-response relationship with intervention duration (4–20 weeks), suggesting that even relatively brief programmes may confer EF benefits. The low heterogeneity (*I*^2^ = 0.0%) supports the construct homogeneity of the pooled estimate, though this should be interpreted cautiously given the limited power of *I*^2^ with small *k*. Preliminary mediation evidence (Wang & Dong, 2026) raises the possibility that EF improvement may serve as an important mediating pathway through which MBI supports broader socio-emotional development, though this requires replication. Future research should prioritise large, pre-registered RCTs with active controls, longitudinal follow-up, and paediatric neuroimaging to strengthen the evidence base and clarify mechanisms.

## Protocol registration

PROSPERO CRD__________________ (registration submitted prior to data extraction).

## Appendix: Coauthor Comment Responses

**Comment 0** (nathanmyli): *Broader EF definitions?* — **Addressed.** Added paragraph in Introduction (Section 1) acknowledging broader constructs (attention, metacognition, behavioral regulation) that neighbour EF but fall outside Diamond’s (2013) tripartite framework, and clarifying why this review retains the strict definition.

**Comment 1** (nathanmyli): *Attention as foundational mechanism?* — **Addressed.** Added paragraph in Introduction citing Posner & Rothbart (2007), Jha et al. (2007), and Lutz et al. (2008) on attention regulation as a mechanism linking mindfulness practice to EF gains.

**Comment 2** (nathanmyli): *Mechanism paragraph needed?* — **Addressed.** Section 4.8 (Mechanistic Considerations) expanded with Wang & Dong (2026) mediation evidence (mindfulness → EF → emotion regulation), tempered with appropriate caveats per Kazdin (2007).

**Comment 3** (Youmans, Scott): *British spellings.* — **No change needed.** The manuscript targets BMJ, which uses British English. Spellings such as “summarised,” “programmes,” and “randomised” are consistent with BMJ house style.

**Comment 4** (Youmans, Scott): *“Expand?” on Study Selection.* — **Addressed.** Section 3.1 now includes the March 2026 update search and expanded exclusion counts (39 excluded at full text with itemised reasons). Section 2.3 details the two-phase, two-reviewer screening protocol.

**Comment 5** (nathanmyli): *Active control discussion?* — **Addressed.** Section 4.10 (Limitations) and Table 1 now explicitly note that 4 of 13 studies used active controls, and that Wang & Dong (2026) used an arts-based active control, strengthening internal validity.

**Comment 6** (nathanmyli): *Age heterogeneity caveat?* — **Addressed.** Section 3.2 now includes a sentence acknowledging the broad developmental range (4–14 years) and directing readers to the subgroup and meta-regression analyses. Section 4.4 discusses overlapping CIs between preschool and school-age subgroups.

**Comment 7** (Youmans, Scott): *“Overall as?” on RoB text.* — **Addressed.** Section 3.4 now reads “eight were rated as overall low risk of bias” and “five as having some concerns,” with explicit per-study attribution and reasoning for each rating level.

**Comment 8** (Youmans, Scott): *RoB domain detail?* — **Addressed.** Section 3.4, paragraph 2, now specifies which studies had concerns in each RoB 2 domain (Domain 1: four quasi-randomised studies; Domain 2: two studies with blinding concerns; Domain 4: one teacher-rated measure).

**Comment 9** (nathanmyli): *Neural/mechanistic evidence from new studies?* **Addressed.** Section 4.8 now discusses Wang & Dong (2026) mediation evidence (EF mediates 53.8% of the mindfulness → emotion regulation pathway), with appropriate caveats noting that this single-study finding requires replication. Paediatric neuroimaging is identified as a research priority.

## Data Availability

This is a systematic review, so we do not have the data to share. The evidence tables in the manuscript have displayed all the data we have extracted.

All data underlying this review are presented in Tables 1–6. The following supplementary materials are available: **Supplementary Table S1**, full electronic search strategies for PubMed and PsycINFO; **Supplementary Table S2**, complete list of excluded studies with reasons; **Supplementary Table S3**, completed PRISMA 2020 checklist with page numbers; **Supplementary File S1**, Python analysis code; **Supplementary File S2**, data extraction spreadsheet. Analysis code and data are available at https://osf.io/[to_be_deposited].

## References

Blair, C., & Razza, R. P. (2007). Relating effortful control, executive function, and false belief understanding to emerging math and literacy ability in kindergarten. Child Development, 78(2), 647–663.

Blakemore, S.-J., & Choudhury, S. (2006). Development of the adolescent brain: Implications for executive function and social cognition. Journal of Child Psychology and Psychiatry, 47(3-4), 296–312.

Borenstein, M., Hedges, L. V., Higgins, J. P. T., & Rothstein, H. R. (2009). Introduction to meta-analysis. Chichester, UK: Wiley.

Chiesa, A., Calati, R., & Serretti, A. (2011). Does mindfulness training improve cognitive abilities? A systematic review of neuropsychological findings. Clinical Psychology Review, 31(3), 449–464.

Cohen, J. (1988). Statistical power analysis for the behavioral sciences (2nd ed.). Hillsdale, NJ: Erlbaum.

Conway, A. R. A., Kane, M. J., Bunting, M. F., Hambrick, D. Z., Wilhelm, O., & Engle, R. W. (2005). Working memory span tasks: A methodological review and user’s guide. Psychonomic Bulletin & Review, 12(5), 769–786.

Cowan, N. (2008). What are the differences between long-term, short-term, and working memory? Progress in Brain Research, 169, 323–338.

DerSimonian, R., & Laird, N. (1986). Meta-analysis in clinical trials. Controlled Clinical Trials, 7(3), 177–188.

Diamond, A. (2013). Executive functions. Annual Review of Psychology, 64, 135–168.

Diamond, A., & Ling, D. S. (2016). Conclusions about interventions, programs, and approaches for improving executive functions that appear justified and those that, despite much hype, do not. Developmental Cognitive Neuroscience, 18, 34–48.

Dunning, D. L., Griffiths, K., Kuyken, W., Crane, C., Foulkes, L., Parker, J., & Dalgleish, T. (2019). Research review: The effects of mindfulness-based interventions on cognition and mental health in children and adolescents—a meta-analysis of randomized controlled trials. Journal of Child Psychology and Psychiatry, 60(3), 244–258.

Duval, S., & Tweedie, R. (2000). Trim and fill: A simple funnel-plot-based method of testing and adjusting for publication bias in meta-analysis. Biometrics, 56(2), 455–463.

Egger, M., Davey Smith, G., Schneider, M., & Minder, C. (1997). Bias in meta-analysis detected by a simple, graphical test. BMJ, 315(7109), 629–634.

Felver, J. C., Tipsord, J. M., Morris, M. J., Racer, K. H., & Dishion, T. J. (2014). The effects of mindfulness-based intervention on children’s attention regulation. Journal of Attention Disorders, 21(10), 872–881.

Flook, L., Goldberg, S. B., Pinger, L., & Davidson, R. J. (2015). Promoting prosocial behavior and self-regulatory skills in preschool children through a mindfulness-based kindness curriculum. Developmental Psychology, 51(1), 44–51.

Flook, L., Smalley, S. L., Kitil, M. J., Galla, B. M., Kaiser-Greenland, S., Locke, J., … Kasari, C. (2010). Effects of mindful awareness practices on executive functions in elementary school children. Journal of Applied School Psychology, 26(1), 70–95.

Flook, L., Goldberg, S. B., Pinger, L., & Davidson, R. J. (2024). The effects of mindfulness training on cognitive control in children: A randomized controlled trial. Applied Developmental Science, 29(2), 141–160.

Gioia, G. A., Isquith, P. K., Guy, S. C., & Kenworthy, L. (2000). Behavior Rating Inventory of Executive Function (BRIEF). Odessa, FL: Psychological Assessment Resources.

Guyatt, G. H., Oxman, A. D., Vist, G. E., Kunz, R., Falck-Ytter, Y., Alonso-Coello, P., & Schunemann, H. J. (2008). GRADE: An emerging consensus on rating quality of evidence and strength of recommendations. BMJ, 336(7650), 924–926.

Haines, B.A., Hong, P.Y., Immel, K.R. et al. The Mindfulness-Based Kindness Curriculum for Preschoolers: An Applied Multi-Site Randomized Control Trial. Mindfulness 14, 2195–2210 (2023). 10.1007/s12671-023-02210-8

Hedges, L. V. (1981). Distribution theory for Glass’s estimator of effect size and related estimators. Journal of Educational Statistics, 6(2), 107–128.

Hölzel, B. K., Lazar, S. W., Gard, T., Schuman-Olivier, Z., Vago, D. R., & Ott, U. (2011). How does mindfulness meditation work? Proposing mechanisms of action from a conceptual and neural perspective. Perspectives on Psychological Science, 6(6), 537–559.

Higgins, J. P. T., Thompson, S. G., Deeks, J. J., & Altman, D. G. (2003). Measuring inconsistency in meta-analyses. BMJ, 327(7414), 557–560.

IntHout, J., Ioannidis, J. P. A., & Borm, G. F. (2014). The Hartung-Knapp-Sidik-Jonkman method for random effects meta-analysis is straightforward and considerably outperforms the standard DerSimonian-Laird method. BMC Medical Research Methodology, 14, 25.

Jha, A. P., Krompinger, J., & Baime, M. J. (2007). Mindfulness training modifies subsystems of attention. *Cognitive, Affective*, & Behavioral Neuroscience, 7(2), 109–119.

Janz, P., Dawe, S., & Wyllie, M. (2019). Mindfulness-based program embedded within the existing curriculum improves executive functioning and behavior in young children: A waitlist controlled trial. Frontiers in Psychology, 10, 2052.

Kabat-Zinn, J. (1994). Wherever you go, there you are: Mindfulness meditation in everyday life. New York: Hyperion.

Kazdin, A. E. (2007). Mediators and mechanisms of change in psychotherapy research. Annual Review of Clinical Psychology, 3, 1–27.

Kander, H., Heininga, V. E., & de Moor, M. H. M. (2024). Effects of mindfulness-based interventions on executive function in children and adolescents: A meta-analysis. Mindfulness, 15, 1–18.

Lutz, A., Slagter, H. A., Dunne, J. D., & Davidson, R. J. (2008). Attention regulation and monitoring in meditation. Trends in Cognitive Sciences, 12(4), 163–169.

Mak, C., Whittingham, K., Cunnington, R., & Boyd, R. N. (2018). Efficacy of mindfulness-based interventions for attention and executive function in children and adolescents—a systematic review. Mindfulness, 9(1), 59–78.

McClelland, M. M., Cameron, C. E., Duncan, R., Bowles, R. P., Acock, A. C., Miao, A., & Pratt, M. E. (2014). Predictors of early growth in academic achievement: The Head-Toes-Knees-Shoulders task. Frontiers in Psychology, 5, 599.

Miyake, A., Friedman, N. P., Emerson, M. J., Witzki, A. H., & Howerter, A. (2000). The unity and diversity of executive functions and their contributions to complex “frontal lobe” tasks: A latent variable analysis. Cognitive Psychology, 41(1), 49–100.

Moffitt, T. E., Arseneault, L., Belsky, D., Dickson, N., Hancox, R. J., Harrington, H., … Caspi, A. (2011). A gradient of childhood self-control predicts health, wealth, and public safety. Proceedings of the National Academy of Sciences, 108(7), 2693–2698.

Page, M. J., McKenzie, J. E., Bossuyt, P. M., Boutron, I., Hoffmann, T. C., Mulrow, C. D., … Moher, D. (2021). The PRISMA 2020 statement: An updated guideline for reporting systematic reviews. BMJ, 372, n71.

Posner, M. I., & Rothbart, M. K. (2007). Research on attention networks as a model for the integration of psychological science. Annual Review of Psychology, 58, 1–23.

Pustejovsky, J. E., & Tipton, E. (2022). Meta-analysis with robust variance estimation: Expanding the range of working models. Prevention Science, 23(3), 425–438.

Parker, A. E., Kupersmidt, J. B., Mathis, E. T., Scull, T. M., & Sims, C. (2014). The impact of mindfulness education on elementary school students: Evaluation of the Master Mind program. Advances in School Mental Health Promotion, 7(3), 184–204.

Quach, D., Mano, K. E. J., & Alexander, K. (2016). A randomized controlled trial examining the effect of mindfulness meditation on working memory capacity in adolescents. Journal of Adolescent Health, 58(5), 489–496.

Ricarte, J. J., Ros, L., Latorre, J. M., & Beltran, M. T. (2015). Mindfulness-based intervention in a rural primary school: Effects on attention, concentration and mood. International Journal of Cognitive Therapy, 8(3), 1–13.

Schonert-Reichl, K. A., & Lawlor, M. S. (2010). The effects of a mindfulness-based education program on pre- and early adolescents’ well-being and social and emotional competence. Mindfulness, 1(3), 137–151.

Schonert-Reichl, K. A., Oberle, E., Lawlor, M. S., Abbott, D., Thomson, K., Oberlander, T. F., & Diamond, A. (2015). Enhancing cognitive and social-emotional development through a simple-to-administer mindfulness-based school program for elementary school children: A randomized controlled trial. Developmental Psychology, 51(1), 52–66.

Sterne, J. A. C., Savovic, J., Page, M. J., Elbers, R. G., Blencowe, N. S., Boutron, I., … Higgins, J. P. T. (2019). RoB 2: A revised tool for assessing risk of bias in randomised trials. BMJ, 366, l4898.

Tang, Y.-Y., Holzel, B. K., & Posner, M. I. (2015). The neuroscience of mindfulness meditation. Nature Reviews Neuroscience, 16(4), 213–225.

Vickery, C. E., & Dorjee, D. (2016). Mindfulness training in primary school decreases negative affect and increases meta-cognition in children. Frontiers in Psychology, 6, 2025.

Viglas, M., & Perlman, M. (2018). Effects of a mindfulness-based program on young children’s self-regulation, prosocial behavior and hyperactivity. Journal of Child and Family Studies, 27(4), 1150–1161.

von Hippel, P. T. (2015). The heterogeneity statistic *I*^2^ can be biased in small meta-analyses. BMC Medical Research Methodology, 15, 35.

Wang, L., & Dong, G. (2026). Mindfulness-based intervention improves executive function and emotional regulation in school-aged children: A randomized controlled trial. Frontiers in Psychology, 17, 1760807. 10.3389/fpsyg.2026.1760807

Zainal, N. H., & Newman, M. G. (2023). Mindfulness-induction and cognition: A systematic review and meta-analysis. Clinical Psychology Review, 102, 102291.

Zeidan, F., Johnson, S. K., Diamond, B. J., David, Z., & Goolkasian, P. (2010). Mindfulness meditation improves cognition: Evidence of brief mental training. Consciousness and Cognition, 19(2), 597–605.

Zelazo, P. D. (2015). Executive function: Reflection, iterative reprocessing, complexity, and the developing brain. Developmental Review, 38, 55–68.

Zelazo, P. D., Forston, J. L., Masten, A. S., & Carlson, S. M. (2018). Mindfulness plus reflection training: Effects on executive function in early childhood. Frontiers in Psychology, 9, 208.

Zenner, C., Herrnleben-Kurz, S., & Walach, H. (2014). Mindfulness-based interventions in schools—a systematic review and meta-analysis. Frontiers in Psychology, 5, 603.

